# Flowdef: An MATLAB Application for Artifact-Reduced Choriocapillaris Flow-Void Quantification

**DOI:** 10.1101/2025.07.05.25330925

**Authors:** Hikmet Yucel, Kholood Janjua, Cem Kesim, Murat Hasanreisoglu, Muhammad Sohail Halim, Mohammed Abdul Saqhlain Shaik, Mohamed I. Ahmed, Carl-Gustav Olsen-Glittenberg, Marieh Esmaeelpour, Victor Chung, Quan Nguyen, Yasir J. Sepah

**Author notes:** **Funding Information** This research received no specific grant from any funding agency in the public, commercial, or not for profit sectors. **Commercial Relationships Disclosures:** None.

## Abstract

**Purpose:** To introduce and validate **Flowdef**, an open□source MATLAB application that automatically excludes drusen□ and vessel□related projection artifacts and enables region□specific quantification of choriocapillaris (CC) flow voids in eyes with age□related macular degeneration (AMD).

**Methods:** Thirty eyes (10 healthy controls, 10 intermediate AMD, 10 dry AMD with geographic atrophy [GA]) underwent 6 × 6 mm OCT□angiography. Flowdef generates exclusion masks for drusen (Otsu threshold × 1.2) and superficial vessels (CLAHE + morphology) and identifies CC flow voids using a “mean ‐ 1 SD” threshold. For GA eyes, an interactive affine registration aligns infrared and OCT en face images to delineate healthy, transition, and atrophic zones. Repeatability (same□day test–retest) and inter□rater reproducibility were assessed with intraclass correlation coefficients (ICC) and coefficients of variation (CV).

**Results:** Flowdef processed all 30 eyes without failure. After artifact exclusion, atrophic zones showed the largest mean flow-void area (1078 µm^2^) while healthy zones exhibited more numerous but smaller voids (mean area 871 µm^2^). Test–retest repeatability was excellent in GA (ICC 0.92–0.99) and control eyes (ICC 0.95–0.99) and variable in intermediate AMD (ICC 0.08-0.80). Inter-rater CV was <10% for most parameters except mean area in intermediate AMD (CV ≈200%).

**Conclusions:** Flowdef provides a robust, freely available solution for artifact□reduced CC flow□void analysis and region□specific GA assessment, addressing key limitations of existing methods.

**Translational Relevance:** By delivering reliable CC metrics with minimal user input, Flowdef can support longitudinal AMD monitoring and accelerate clinical research on emerging therapies.

## INTRODUCTION

Choriocapillaris flow voids are emerging biomarkers in retinal disease [1–3]. Because the CC supplies the outer retina, accurate measurement of its perfusion is critical for understanding age□ related macular degeneration (AMD) progression and response to therapy.

Existing OCTA□based approaches face two persistent obstacles: (i) projection and shadow artifacts from overlying vessels and drusen that masquerade as CC non□perfusion, and (ii) lack of region□specific analysis within the same eye, particularly across healthy retina, transition zone (TZ), and geographic atrophy (GA).

We therefore developed **Flowdef**, a user□friendly, open□source MATLAB application that (i) excludes artifact□prone pixels before thresholding, (ii) implements interactive registration so that GA, TZ, and healthy areas can be quantified separately, and (iii) outputs standardized metrics (total, mean, percent area; void counts by size categories) suitable for longitudinal or multi□center studies.

Here we describe Flowdef’s algorithms and present validation data from 60 eyes spanning the AMD severity spectrum.

## METHODS

### Study Population

This retrospective study analyzed 30 eyes from 30 subjects imaged under an IRB-approved protocol with written informed consent. The study cohort comprised 10 healthy controls, 10 eyes with intermediate AMD (Age-Related Eye Disease Study [AREDS] stage 3; large drusen, no GA or CNV), and 10 eyes with dry AMD with GA (confirmed on fundus AF and OCT). Mean ages were 63.8 ± 4.8, 73.8 ± 10.8, and 81.3 ± 3.2 years for controls, intermediate AMD, and GA groups, respectively. Controls were predominantly female, intermediate AMD patients were all male, and GA patients were all female. The majority of participants across all groups were White. All imaging and measurements were obtained from screening examinations.

### Image Acquisition

All subjects underwent 6 × 6 mm macular OCT□angiography on an RTvue□XR Avanti SD□OCTA (Optovue, Fremont CA). Only scans with signal□quality index ≥7 and free of gross motion artifacts were included.

### Description of the application

Flowdef app is a Matlab application with a detailed graphical user interface (GUI) as shown in Figure 1. With this app, the user can perform the following functions:

**Figure 1.**
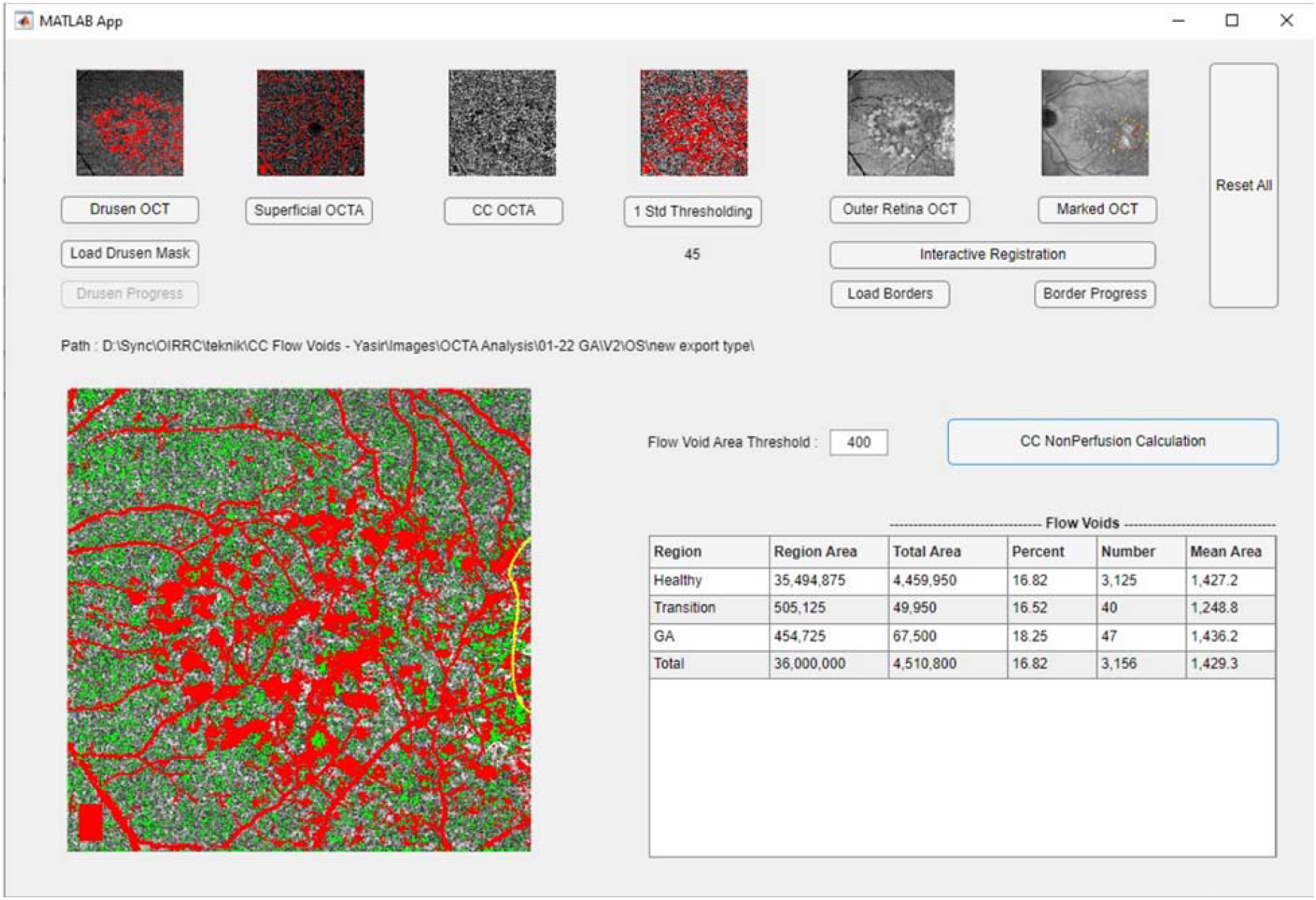
Graphics User Interface (GUI) of the Flowdef application.

- Visualize flow voids in the CC layer of OCTA
- Automatically generate masks for drusen and vessels on using FAF, IR and OCT en face images
- Manually edit masks with any third party app, and save the masks for later use,
- Register different OCT images to map the manually annotated borders,
- Determine the area of regions between the borders
- Calculate the total area, percentage, number and mean area of flow voids for each region
- Track the progression of drusen regions over time and changes in the areas of various lesions such as scotoma, transition zone and normal retina.

### Image Acquisition and Quality Control

OCTA images were acquired using the RTvue-XR Avanti spectral domain-OCTA system (Optovue, Inc, Fremont, CA). All subjects underwent 6×6 mm en face OCTA analysis with scans matched for signal quality index.

### Segmentation Methodology

Before processing the CC flow void, the Flowdef app generates masks for shadows cast by drusen and large vessels following the algorithms that are described below:

### Artifact□Exclusion Algorithms

#### Drusen segmentation

A special slab on which drusen can be seen clearly is extracted from the OCT en face image. The app first reads the image and converts it to grayscale, then it adjusts the image intensity values. This process saturates the bottom 1% and the top 1% of all pixel values and increases the contrast of the image. Since drusens are brighter than the other regions, global thresholding is used to detect the drusen in the literature. In this study, global thresholding with Otsu method is used to determine a threshold and 1.2 times of it is used as a threshold for segmentation [5]. The 1.2 value was determined empirically as an optimum value. The detailed implementation is provided in Supplementary Material 1.

### Vessel segmentation Algorithm

The algorithm starts with reading the image and converting it to grayscale. Generally, the brightness is not flat in some superficial OCTA images, especially taken from SS devices. Contrast limited adaptive histogram equalization (CLAHE) algorithm is applied to the image first to solve this issue. The vessels in the superficial OCTA image are generally brighter than the other regions. However, there are other brighter regions in the image, therefore, global thresholding remains insufficient for segmentation.

Since the vessels are tubular shapes, shape information must be used as well to differentiate the vessels from other bright spots. This algorithm uses a detail level threshold value to decide how much thin vessels will be segmented. A higher value means thinner vessels will be segmented. Global thresholding with the Otsu method Reference is used to determine a threshold. 1.2 times of it for SD devices and 1.5 times of it for SS devices is used as a threshold for binarization.

The app gets the skeleton of the binarized image, but it omits the branches less than 100 pixels. Thus, unconnected bright spots which are not vessels don’t have a skeleton. This skeleton is used to fill the inverse of the binarized image. Filling is started from the skeleton pixels and spreads to the connected pixels. By this way, vessels are filled whereas unconnected bright spots cannot be, and the result is the bright spots only. Finally, the vessels are obtained by taking the difference between the binarized image and the bright spots. The union of drusen + vessel masks form the total exclusion mask.

### Flow Void Detection Methodology

Flow void detection was performed using the “1 standard deviation below mean” threshold method. The average pixel intensity and standard deviation of the CC OCTA image were calculated after excluding pixels in the total exclusion mask. The nonperfusion threshold was determined by subtracting the standard deviation of pixel brightness from the mean brightness value. Pixels with intensity values below this threshold were classified as flow deficits within the CC slab. The detailed flow void detection algorithm implementation is provided in Supplementary Material 1.

### Region□Specific GA Analysis

For GA eyes, borders of GA and TZ were manually drawn on the infrared image; 8–10 vessel bifurcations were paired to the OCT en face slab to compute an affine transformation (Figure 6). Masks for GA, TZ, healthy, and combined regions were generated automatically after registration.

To facilitate accurate analysis, we map the delineated borders from the Infrared (IR) image to the corresponding enface slab of the Optical Coherence Tomography (OCT). This mapping is achieved through an interactive registration process. Users identify and mark corresponding points on both images, typically choosing intersections or bifurcations of vessels as these are reliable landmarks.

Following this, our application computes an affine transformation based on these point pairs and applies this transformation to warp the IR image accordingly.

The core of the analysis methodology computes the sizes of the flow deficit areas using a flood-fill algorithm (Ref) that searches for deficit pixels. The employed algorithm initiates at the top left corner of the image, searching for the nearest deficit pixel. Upon finding one, it launches a flood-fill operation from this pixel, effectively covering the entire deficit area.

### Validation of Repeatability and Reproducibility

For repeatability testing, two consecutive OCTA scans were obtained from the same eye with a 5 minute interval between acquisitions, and flow void analysis was performed by the same observer. The coefficient of variation (CV) between paired measurements was calculated for each flow void parameter, and intraclass correlation coefficient (ICC) values were determined to quantify test-retest reliability.

To evaluate inter-rater reproducibility of Flowdef measurements, two independent raters analyzed the same set of OCTA images, making manual adjustments to the algorithm-generated vessel and drusen masks where deemed necessary. Coefficient of variation (CV) was calculated for each flow void parameter to quantify the relative variability between raters, with CV values below 10% considered to indicate good measurement reproducibility. Additionally, we compared both raters’ manually adjusted measurements to the fully automated results to assess the impact of manual adjustments on flow void quantification. These analyses were stratified by cohort and, for GA eyes, by anatomical regions (healthy area, transition zone, atrophic area, and full image) to identify parameters and regions with the highest measurement consistency.

## RESULTS

### Segmentation Performance

When applying our drusen segmentation algorithm to OCTA en face images, we obtained distinct masks that highlighted drusen areas as shown in Figure 5A. The algorithm effectively identified the brighter areas corresponding to drusen, providing a basis for their exclusion in subsequent flow void analysis.The vessel segmentation process successfully identified the vascular structures in the superficial OCTA images, as demonstrated in Figure 5B. Our approach distinguished vessels from other bright areas in the image by leveraging both brightness and structural information, resulting in accurate vessel masks.

### Combined Exclusion Mask and Flow Void Detection

In the GUI of the app, after loading drusen OCT, superficial OCTA and CC OCTA images, the “1 Std Thresholding” button is enabled. Clicking this button, first combines the drusen and vessel mask to obtain a total exclusion mask and displays it on the CC OCTA image as in Figure 5C.

The average pixel intensity and the standard deviation of pixel intensity of the CC OCTA image are calculated after excluding the pixels in the total exclusion mask. Then the threshold is calculated and displayed under the button as in Figure 5D. Pixels with brightness levels below the threshold are identified as flow deficits within the slab. The algorithm computed the sizes of the flow deficit areas after the exclusion of areas covered by drusen and vessel mask. For subjects with GA, the analysis was further divided into GA lesion, TZ and healthy retina.

After calculating the nonperfusion threshold, the nonperfusion areas are classified according to this flow void area threshold. The app calculates the total area, mean area, percentage, and number of flow voids for each region.

### Image Registration Results

The image registration process successfully mapped the annotated borders from IR images to OCT en face images. Figure 6 shows the results of this registration process. Figure 6A demonstrates the interface for matching corresponding points between the two imaging modalities. Figure 6B shows an infrared image with manually annotated borders of geographic atrophy (red) and transitional zone (yellow).

To verify the precision of this transformation, the application overlays the warped IR image onto the OCT slab, presenting it in a checkerboard pattern along with the transformed borders for visual inspection. Users can assess the transformation’s accuracy by examining the continuity of the vessels within the checkerboard display, as shown in Figures 6C and 6D.

### Flow Void Analysis Across Different Regions

The Flowdef application successfully processed all 30 eyes and generated quantitative flow void metrics for each anatomical region. The extent of the filled area is determined by subtracting the flood-filled image from the original, with the size of the deficit quantified by the count of filled pixels as shown in Figure 2. These measurements are stored in an array. The outcome is an array detailing the sizes of each flow deficit area. These results are then presented both visually and numerically.

**Figure 2.**
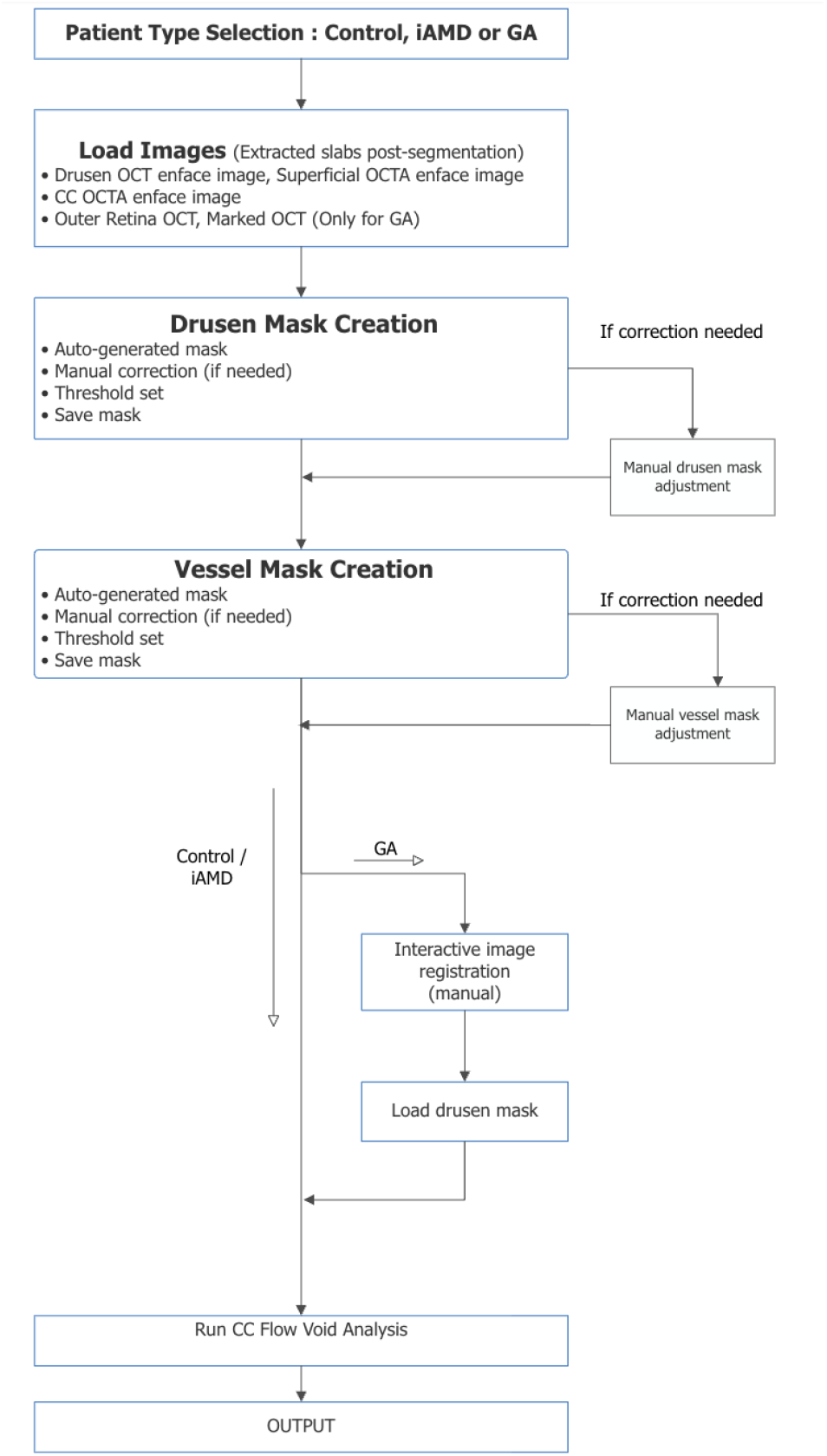
Flowdef application workflow showing the complete process from patient type selection through flow void analysis.

**Figure 3.**
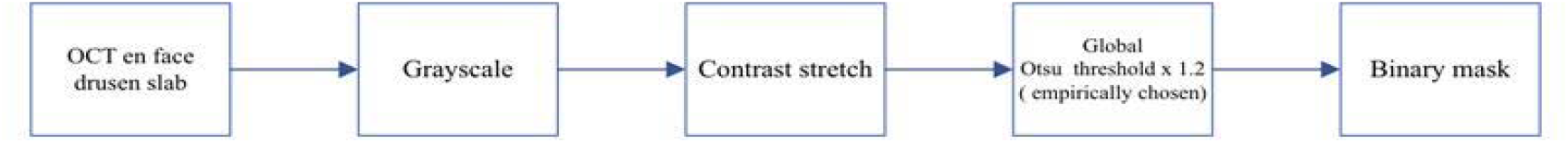
Image processing workflow for automated drusen segmentation from OCT en face images.

**Figure 4.**
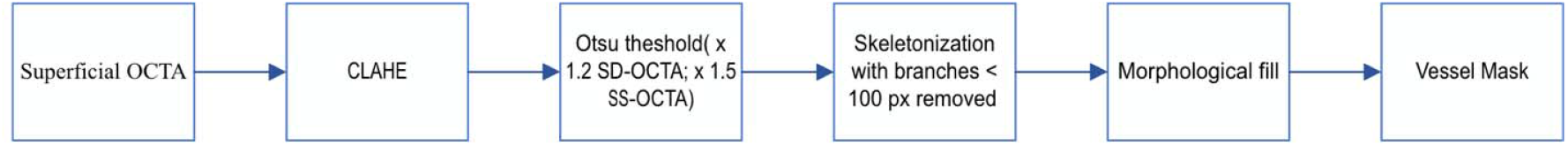
OCTA vessel segmentation workflow for superficial capillary plexus analysis.

**Figure 5.**
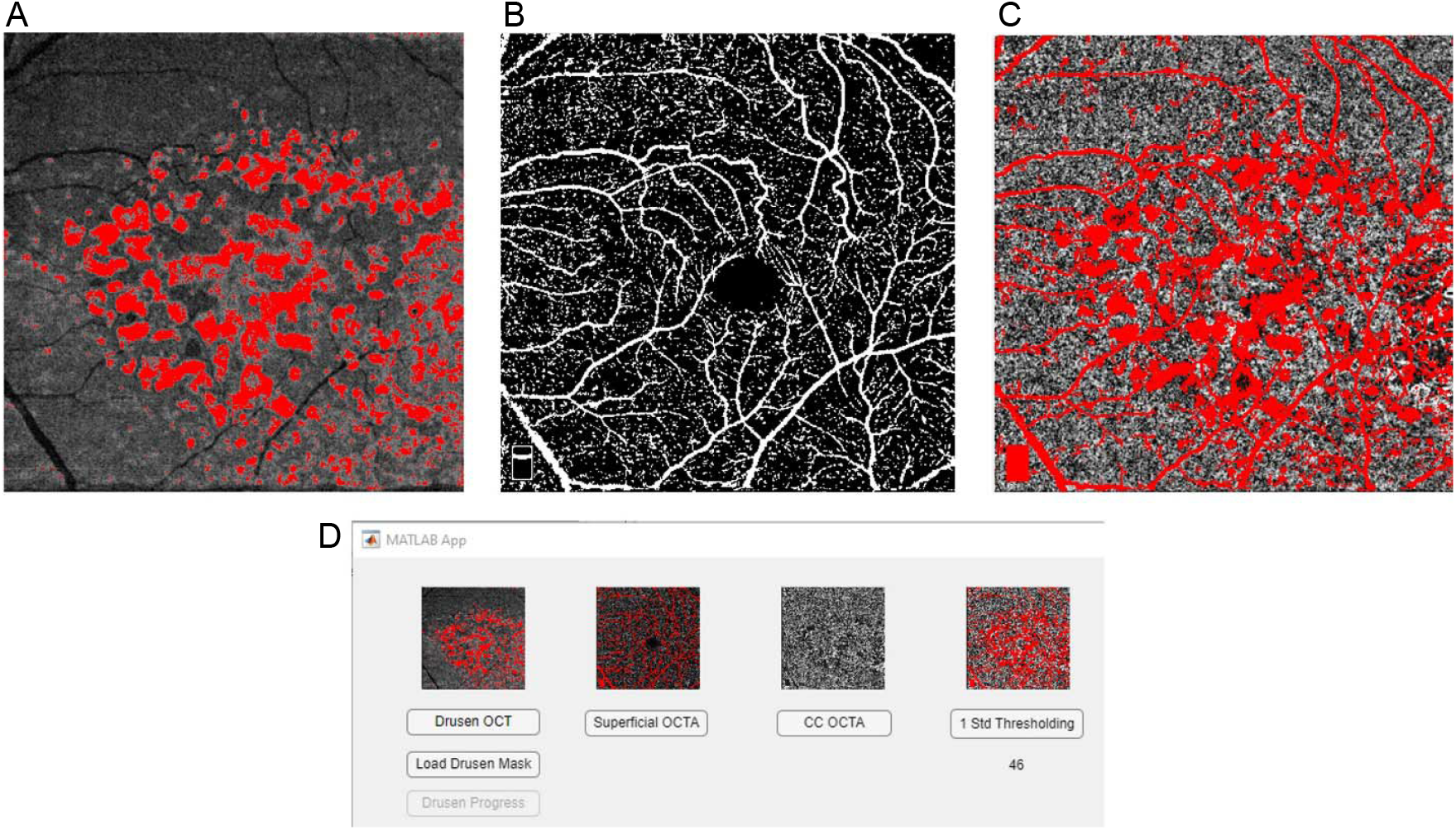
Segmentation processing in the Flowdef app. A: Drusen mask generated by segmentation process is optionally loaded over the en face OCT image. B: Binarized superficial OCTA image. C: Total exclusion mask on CC OCTA image. D: Standard thresholding process of the application.

**Figure 6.**
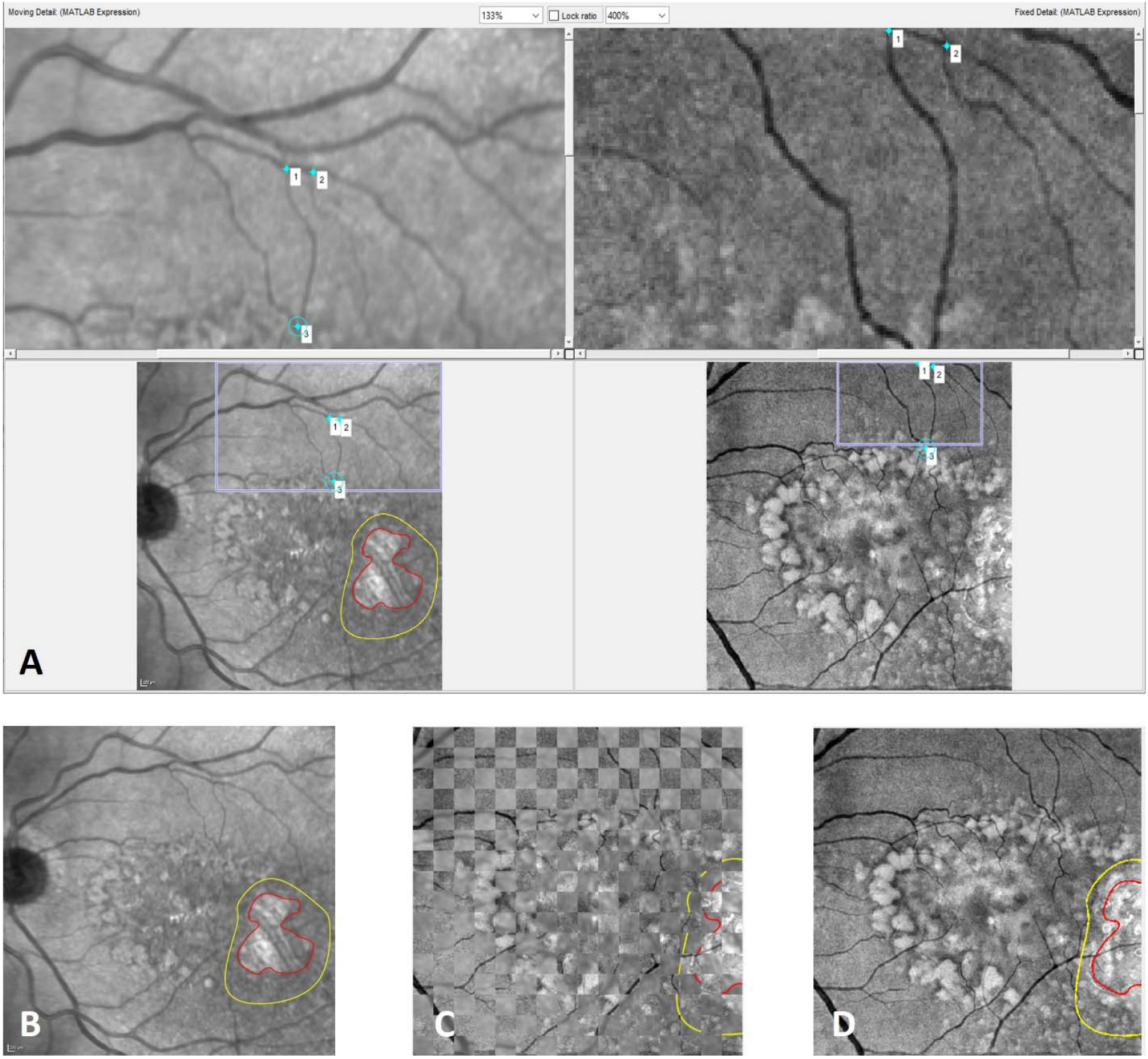
Image transposition of the Flowdef app. A: Infrared and OCT en face image matching process in the custom-made graphical user interface. B: Infrared image of a patient with annotated borders of geographic atrophy (red) and transitional zone (yellow). C and D: Checkerboard display of the infrared and en face OCT image merging process of the application.

Our preliminary analysis across different regions (N = 30 eyes; 10 healthy controls, 10 intermediate AMD, and 10 geographic atrophy patients) is summarized in Table 1. The table shows the distribution of flow voids across different size categories (200µm^2^, 400µm^2^, 600µm^2^, ≥800µm^2^) for each region.

**Table 1:** Flow void characteristics across different regions and pathologies.

| Cohort | Region | Mean area ($\mu\text{m}^2$) | Count 200$\mu\text{m}^2$ | Count 400$\mu\text{m}^2$ | Count 600$\mu\text{m}^2$ | Count $\geq 800\mu\text{m}^2$ |
| --- | --- | --- | --- | --- | --- | --- |
| Control | Full image | 855 | 2,436 | 1,117 | 632.2 | 1,745 |
| iAMD | Full image | 952 | 2,172 | 962.5 | 535.9 | 1,567 |
| GA | Full image | 904.42 | 2,168 | 911.16 | 527.16 | 1,498 |
|  | Healthy area | 871.18 | 1,788 | 752.16 | 435.8 | 1,207 |
|  | Transition zone | 889.02 | 218.5 | 90.5 | 52.3 | 147.3 |
|  | Atrophic area | 1078.4 | 200.3 | 83.5 | 47.1 | 150.16 |

The atrophic areas showed the highest mean flow void area (1078.4 μm^2^) compared to healthy areas (871.18 μm^2^) and transition zones (889.02 μm^2^). However, the absolute number of flow voids was highest in healthy areas across all size categories.

Notably, the flow signal within the geographic atrophy areas showed evidence of disturbance by excessive hyperintense signals from deeper choroidal vessels, which is likely due to window defects of RPE and the disappearance of choriocapillaris beneath the atrophy.

### Reproducibility and Repeatability Results

The Flowdef application demonstrated variable test-retest repeatability across different cohorts and parameters (Table 2). GA eyes showed the highest consistency with ICC values ranging from 0.92 to 0.99 across all measurements. Control eyes demonstrated good to excellent repeatability (ICC: 0.95-0.99) for most parameters. iAMD eyes showed more variable reproducibility, with ICC values ranging from 0.08 to 0.8, reflecting the heterogeneous nature of disease presentation in this cohort.

**Table 2:** Intraclass Correlation Coefficient (ICC) for Test-Retest Repeatability - ICC (95% CI)

| Cohort | Total Area | Mean Area | Percent Area | Number 200$\mu\text{m}^2$ | Number 400$\mu\text{m}^2$ | Number 600$\mu\text{m}^2$ | Number $\geq 800\mu\text{m}^2$ | Number Total |
| --- | --- | --- | --- | --- | --- | --- | --- | --- |
| Control | 0.96<br>(0.5-0.99) | 0.97<br>(0.9-0.99) | 0.972<br>(0.89-0.99) | 0.95<br>(0.17-0.99) | 0.96<br>(0.67-0.99) | 0.99<br>(0.88-0.99) | 0.98<br>(0.61-0.99) | 0.96<br>(0.02-0.99) |
| iAMD | 0.65 | 0.682 | 0.08 | 0.530( | 0.536 | 0.72 | 0.8 | 0.51 |
|  | (0.12-0.89) | (0.13-0.91) | (0.5-0.6) | 0.16-0.86) | (0.14-0.86) | (0.19-0.92) | (0.3-0.9) | (0.06-0.84) |
| GA | 0.92<br>(0.89-0.99) | 0.99<br>(0.96-0.99) | 0.99<br>(0.98-1.0) | 0.99<br>(0.98-1.0) | 0.99<br>(0.98-1.0) | 0.98<br>(0.97-0.99) | 0.99<br>(0.98-1.0) | 0.99<br>(0.98-1.0) |

Inter-rater reproducibility assessment revealed variation across different cohorts and measurement parameters (Table 3). Control eyes demonstrated consistent inter-rater agreement with coefficient of variation (CV) values generally below 10% for most parameters, except for mean area (9.8%). iAMD eyes showed higher variability, particularly for mean area (CV = 200.7%) and percent area (CV = 50.4%), while other parameters remained more stable (CV range: 3.2-19.8%).

**Table 3:** Coefficient of Variation (Mean ± SD) for Inter-rater Reproducibility by Cohort.

| Cohort | Total Area (%) | Mean Area (%) | Percent Area (%) | Number 200μm<sup>2</sup> (%) | Number 400μm<sup>2</sup> (%) | Number 600μm<sup>2</sup> (%) | Number ≥800μm<sup>2</sup> (%) | Number Total (%) |
| --- | --- | --- | --- | --- | --- | --- | --- | --- |
| Control | 6.8 | 9.8 | 5.2 | 5.0 | 2.8 | 4.4 | 7.9 | 7.8 |
| iAMD | 3.2 | 200.7 | 50.4 | 19.8 | 7.4 | 6.6 | 10.3 | 8.8 |
| GA | 7.8 | 16.5 | 72.3 | 13.5 | 11.3 | 9.9 | 8.8 | 7.4 |
| Full Image | 11.1 | 15.0 | 11.9 | 11.3 | 9.9 | 8.8 | 7.4 | 6.9 |
| Healthy Area | 19.4 | 13.9 | 10.5 | 12.4 | 11.2 | 14.6 | 15.8 | 12.0 |
| Transition Zone | 67.6 | 22.7 | 17.5 | 53.5 | 59.0 | 64.1 | 65.0 | 56.5 |
| Atrophic Area | 15.8 | 15.1 | 10.8 | 9.3 | 6.6 | 9.3 | 12.3 | 7.1 |

Within GA eyes, region-specific analysis revealed differences in measurement reproducibility. Transition zones showed the highest variability across all parameters (CV range: 17.5-67.6%), reflecting the challenging nature of accurately delineating these borderland areas. Healthy areas within GA eyes demonstrated moderate reproducibility (CV range: 10.5-19.4%), while atrophic areas showed generally acceptable reproducibility (CV range: 6.6-15.8%) for most measurements.

## DISCUSSION

This study introduces **Flowdef**, an open□source MATLAB tool that tackles two perennial obstacles in choriocapillaris (CC) OCTA analysis; projection/shadow artifacts and region□specific quantification inside the same eye, while maintaining high repeatability and acceptable inter□rater agreement.

1. After artifact exclusion, **mean flow**□**void area and void**□**size distribution mirrored the clinical spectrum of AMD**: small, numerous voids in healthy retina; intermediate values in iAMD; and large confluent voids inside GA.
2. **Repeatability was excellent in GA and control eyes** (ICC > 0.95) and lower in iAMD, where drusen heterogeneity amplifies segmentation□threshold noise.
3. **Manual mask editing rarely altered results** except in iAMD, implying the automated pipeline is robust for most clinical scenarios.

### Comparison With Previous Work

The choriocapillaris is a crucial vascular network that provides nutrients and oxygen to the outer retina, including the retinal pigment epithelium (RPE) and photoreceptors. Disruptions in choriocapillaris flow are implicated in several retinal diseases, most notably age-related macular degeneration (AMD). AMD is a leading cause of vision loss among the elderly and manifests in two primary forms: non-exudative (dry) and exudative (wet) AMD. In dry AMD, the accumulation of drusen and the subsequent atrophy of the RPE lead to geographic atrophy (GA), while wet AMD is characterized by choroidal neovascularization (CNV) and leakage of blood and fluid. The integrity of the choriocapillaris is essential for maintaining retinal health and function, and any compromise can lead to significant visual impairment [5,6].

Several studies have focused on visualizing and quantifying choriocapillaris flow voids using optical coherence tomography angiography (OCTA), particularly in advanced stages of AMD, including exudative neovascular AMD and geographic atrophy. Jia et al. highlighted the potential of OCTA to detect choriocapillaris flow voids in AMD patients, associating these voids with areas of ischemia and disease severity [7]. However, significant setbacks remain due to imaging artifacts. Motion artifacts, improper segmentation, and projection artifacts are prevalent issues that compromise the reliability of OCTA images. These artifacts can obscure true choriocapillaris flow deficits, complicating the differentiation between actual pathological changes and imaging errors. Notable studies addressing these issues include Shi et al., who investigated choriocapillaris flow deficits in advanced AMD, and Rinella et al., who explored the relationship between choriocapillaris flow voids and GA [8,9]. Additionally, Nassisi et al. provided insights into the quantification of choriocapillaris flow deficits in neovascular AMD, highlighting the need for improved imaging techniques [10].

To address the challenges posed by imaging artifacts, various studies have proposed methods to reduce or eliminate these artifacts in OCTA. Chu et al. developed an automated method for quantifying choriocapillaris flow deficits, utilizing advanced image processing algorithms to minimize the impact of artifacts and enhance the clarity of OCTA images [11]. Another significant advancement is the use of swept-source OCTA (SS-OCTA), which offers deeper penetration and faster acquisition speeds, reducing motion artifacts and improving image quality. Scharf et al. demonstrated that SS-OCTA, combined with enhanced image processing techniques, significantly improved the accuracy of choriocapillaris imaging by reducing artifact-induced distortions [12]. Other notable studies include reducing artifactual flow deficits using advanced methods, reducing projection artifacts, and implementing image compensation strategies to reduce hyperreflective signals [13-15]. In the current study, instead of the aforementioned strategies which have the risk to put a level of bias on the interpretation of flow in the artifact regions, we directly excluded the areas that are affected by the projection and shadowing artifacts that are due to retinal vessels longer than a predefined 100-pixel length which roughly corresponds to 1.5 mm length.

A critical issue in interpreting OCTA images is whether shadows cast by drusen represent actual drusen-related flow voids or are merely artifacts. Drusen, which are extracellular deposits that accumulate between the RPE and Bruch’s membrane, can obstruct the imaging beam, creating shadows that complicate the interpretation of choriocapillaris flow. Some researchers argue that these shadows are artifacts resulting from the physical blockage by drusen. However, studies like those by Spaide and Curcio suggest that drusen can indeed cause localized disruptions in choriocapillaris perfusion, indicating that these shadows may correspond to true reductions in blood flow [16]. Thus, distinguishing between artifact-induced shadows and genuine flow voids is essential for accurate assessment and monitoring of choriocapillaris health in AMD patients, which was found based on our image processing method that enabled the users to evaluate flow parameters of drusen-free areas separately.

In cases of AMD with geographic atrophy (GA), hyperreflective flow signals in OCTA images pose a significant challenge. These signals, often caused by high reflectivity from underlying tissues, can obscure true choriocapillaris flow deficits, making it difficult to accurately assess the extent of atrophy and perfusion. Geographic atrophy, characterized by extensive RPE and photoreceptor loss, requires precise visualization of choriocapillaris flow to monitor disease progression and evaluate therapeutic interventions. Reducing hyperreflective flow signals is therefore crucial to enhance the clarity and accuracy of OCTA images, allowing for better diagnosis and management of GA in AMD patients [17,18]. Improved imaging techniques can lead to better outcomes by providing clearer images that help in the precise assessment of the choriocapillaris, thereby aiding in the development of targeted treatments. In the current study, we focused on the transitional zone that includes the border of the geographic atrophy and the healthy outer retina rather than the geographic atrophy itself to avoid signal abnormalities that could be induced by the lesion. As shown in the Results section, flow signals within the geographic atrophy areas are heavily disturbed by an excessive hyperintense signal from deeper choroidal vessels which is due to window defect of RPE and the disappearance of choriocapillaris beneath the atrophy, leading into hyper transmission of flow signal from Haller and Sattler layers of the choroid.

### Biological Interpretation

The high void counts but small areas in healthy retina likely reflect the physiological CC mosaic, whereas the shift toward fewer, larger voids in GA suggests a **coalescence model of capillary loss**. The iAMD variability may signify an intermediate, unstable state in which local ischemia fluctuates around drusen even when outer□retinal structure appears preserved.

### Clinical and Research Implications

Because Flowdef outputs standardized metrics stratified by anatomical region, it can: (i) serve as a **biomarker endpoint** in trials testing complement inhibitors or gene therapy for GA; (ii) help identify iAMD eyes with rapid CC deterioration that may benefit from earlier intervention; and (iii) provide a common platform for cross□center data pooling, accelerating biomarker qualification.

### Limitations and Future Directions

The modest sample limits generalizability, and all images were acquired on a single SD□OCTA device. **Multi**□**vendor validation** is underway, and we are integrating a U□Net–based GA/TZ auto□segmentation to eliminate manual border tracing. Finally, hyper□transmission artifacts inside GA remain problematic; future versions will incorporate the attenuation□compensation strategy of Wang et al.^15 as an optional module.

## CONCLUSION

Flowdef delivers artifact□reduced, region□specific CC flow□void metrics with repeatability comparable to, or better than, published OCTA methods, paving the way for reliable longitudinal monitoring of AMD and rapid evaluation of emerging therapies.

## Data Availability

All data produced in the present study are available upon reasonable request to the authors.

## SUPPLEMENTARY MATERIAL 1 - FLOWDEF APPLICATION CODE

### Drusen Segmentation Algorithm

% read the image and convert it to grayscale app.s.drusenOct=rgb2gray(imread([app.s.path,file]));

% calculate the mean and use it as a threshold

thr = 1.2*graythresh(imadjust(app.s.drusenOct));

% binarize the image according to the threshold app.s.drusenMask=imbinarize(app.s.drusenOct,thr);

### Vessel Segmentation Algorithm

#### Initial Image Processing

% read the image and convert it to grayscale app.s.superfOcta=rgb2gray(imread([app.s.path,file]));

% calculate the graylevel threshold

thr = graythresh(adapthisteq(app.s.superfOcta));

if app.s.superfOctaSwept==true

thr=thr*1.5;

else

thr=thr*1.2;

end

% binarize inpImg according to the thr. Since vessels are

% brighter, they are binarized as 1.

tempImg=imbinarize(inpImg,thr);

#### Bright Spot Removal

% get skeleton of tempImg with rejecting branches less than 100px.

% This rejects most of the unconnected pixels which are bright spots. tempImgSkel=bwskel(tempImg,’MinBranchLength’,100);

% get coordinates of the skeleton pixels

[row,col] = find(tempImgSkel);

% invert the binarized image. So, the vessels are 0 now.

tempImgInv=~tempImg;

% fill the vessels starting from the skeleton. Here, all pixels

% connected to skeleton are filled with 1. So, only the unconnected

% bright spots’ pixels are left 0.

tempImgInvFilled=imfill(tempImgInv,[row,col],8);

% get the vessels by subtraction and assign it as

% outImg. Since the only vessels become white by

% filling and the rest is the same, only vessel pixels are 1

% and the rest is 0

outImg=tempImgInvFilled-tempImgInv;

### Flow Void Detection Algorithm

#### Threshold Calculation

% calculate mean of ccOcta except excludeMask

meanOcta=round(mean(double(app.s.ccOcta(~app.s.excludeMask))));

% calculate mean of ccOcta except excludeMask

stdOcta=round(std(double(app.s.ccOcta(~app.s.excludeMask))));

% calculate stdBelowMean

app.s.stdBelowMean=meanOcta-stdOcta;

#### Flow Void Area Calculation

% invert nonPerfused, so nonPerfused pixels are 0 now

nonPerfusIng=~nonPerfused;

% set excludeMask pixels 1 on nonPerfInv

nonPerfInv(excludeMask)=true;

% create array for areas of filled regions

filledPix=[];

% row loop

for row=1:400

% column loop

for col=1:400

% if pixel is nonperfused

if nonPerfInv(row,col)==false

% fill starting from that pixel

nonPerfInvNew=imfill(nonPerfInv,[row,col],8);

% get the filled region by substraction

filled = nonPerfInvNew - nonPerfInv;

% add the area of the filled region to filledPix array

filledPix=[filledPix;sum(sum(filled))];

end

end

end

% update nonPerfInv with the filled one.

nonPerfInv=nonPerfInvNew;

% total area of nonmasked regions totalArea=sum(sum(~excludeMask))*px2Area;

% get values bigger than app.s.thr

thrFilledPix=filledPix(filledPix*px2Area > thr);

% the total area of regions bigger than app.s.thr

thrArea=sum(thrFilledPix)*px2Area;

% area ratio of nonperfused to total

ratio=100*thrArea / totalArea;

% average area of regions bigger than app.s.thr meanThrArea=mean(thrFilledPix)*px2Area;

## Notes

### Competing Interest Statement

The authors have declared no competing interest.

### Author Declarations

This study was approved by the institutional review board of Stanford University (IRB no. 68008) and adhered to the Declaration of Helsinki. Informed consent was obtained from enrolled subjects, and their protected health information was de-identified before storage in password-protected browsers accessible to the immediate study team only.

